# Safety of RSV Vaccine among Pregnant Individuals: A Real-World Pharmacovigilance Study Using Vaccine Adverse Event Reporting System

**DOI:** 10.1101/2024.04.19.24306090

**Authors:** Abdallah Alami, Santiago Perez-Lloret, Donald R. Mattison

**Affiliations:** School of Epidemiology and Public Health, Faculty of Medicine, University of Ottawa, Ottawa, Ontario, Canada; Consejo Nacional de Investigaciones Científicas y Técnicas (CONICET), Buenos Aires, Argentina; Observatorio de Salud Pública, Pontificia Universidad Católica Argentina,Buenos Aires, Argentina; Department of Physiology, Faculty of Medicine, University of Buenos Aires,Buenos Aires, Argentina; Risk Sciences International, Ottawa, Canada

**Author notes:** Corresponding Author: Abdallah Alami, School of Epidemiology and Public Health, University of Ottawa, Ottawa, ON K1G 5Z3, Canada.

## Abstract

**Objectives:** To describe the post-marketing safety of RSVPreF among pregnant individuals.

**Design:** This case series study analyzed adverse event (AE) reports submitted to the U.S. Food and Drug Administration’s Vaccine Adverse Event Reporting System (VAERS) database following RSVPreF immunization from September 1, 2023, to February 23, 2024.

**Setting:** VAERS, as a national spontaneous vaccine safety surveillance system, provides insights into the safety profile of the RSVPreF vaccine in a real-world setting.

**Participants:** Surveillance data included all AE reports submitted to VAERS for pregnant individuals following vaccination.

**Exposure:** Receipt of RSVPreF vaccine among pregnant individuals in the U.S.

**Primary and secondary outcome measures:** Descriptive statistics assessed all AE reports with RSVPreF, including frequency, gestational age at vaccination, time to AE onset, and serious report proportions. The Bayesian Confidence Propagation Neural Network (BCPNN) was utilized, estimating the information component (IC) to identify disproportionate reporting of RSVPreF–event pairs.

**Results:** VAERS received 77 reports pertained to RSVPreF vaccination in pregnant individuals, with 42 (54.55%) classified as serious. The most reported non-pregnancy-specific AEs were headache, injection site erythema, and injection site pain. Preterm birth was the most frequently reported pregnancy-specific AE, followed by preterm premature rupture of membranes, cesarean section, cervical dilatation, and hemorrhage during pregnancy. The median time from immunization to reported preterm birth was 3 days, with two-thirds of cases within a week. Disproportionality analysis indicated a significant signal for various AEs, particularly highlighting preterm birth with an IC of 2.18 (95%CI, 1.54-2.63), suggesting that reports of preterm birth associated with RSVPreF vaccination occurred more frequently than statistically expected.

**Conclusions:** While reported AEs were generally consistent with the safety profile observed in prelicensure studies, this study highlights ongoing concern about preterm birth among pregnant individuals following RSVPreF vaccination. Comprehensive longitudinal follow-up, including prospective pregnancy registries and infant follow-up studies is urgently required.

## INTRODUCTION

Pregnant individuals and their newborn infants face an increased risk from vaccine-preventable diseases and adverse outcomes ^1,2^. Vaccines such as those against influenza, pertussis, and COVID-19 are hence recommended, not only to protect mothers but also to confer immunity to their infants, reducing the risk of severe illnesses. Among the various pathogens of concern, Respiratory Syncytial Virus (RSV) represents a substantial burden for infants under 6 months of age ^3^, accounting for around 1.4 million hospitalizations and 45,700 deaths globally each year ^4^. In the U.S., RSV infections stands as the leading cause of infant hospitalizations among those younger than 6 months ^5^, highlighting the need for an effective intervention to reduce the burden of the disease ^6^, all while weighing potential benefits and risks ^3,5^. In response, the Food and Drug Administration (FDA) has recently approved two agents for protecting infants against severe RSV illness: nirsevimab, a long-acting monoclonal antibody developed for newborns ^7^, and RSVPreF (Abrysvo, Pfizer), a novel RSV vaccine formulated with the prefusion F protein, approved for pregnant individuals ^8^.

In efforts to protect newborns against severe RSV illnesses, GSK and Pfizer have both conducted clinical trials on RSV vaccines for pregnant individuals, with the goal of transferring immunity to their infants ^9^. GSK prematurely halted its trial due to concerns of an elevated risk of preterm birth ^10^ – a condition defined as birth occurring before 37 completed weeks of gestation ^11^.

Similarly, Pfizer’s phase 3 trial reported a numerical, though not statistically significant, rise in preterm births ^12^, capturing all known pathways to preterm delivery, including premature rupture of membranes, preterm labor, and provider-induced preterm birth ^5^. However, the varied time to onset from vaccination to preterm birth, spanning weeks to months, has further challenged the assessment of a direct causal relationship ^5,8^.

Amidst persistent concerns about the potential association between RSV vaccines and an increased risk of preterm birth, the FDA approved RSVPreF and labeled this potential risk as a warning ^8^, while restricting vaccine use to between 32 and 36 weeks of gestation. This intended to lower the likelihood of vaccine-related preterm births ^8,13^.

The uncertainty surrounding the observed safety signal of preterm birth post-RSV vaccination— as a true risk or coincidental—persists due to insufficient data to conclusively establish or refute a causal relationship with the vaccine ^5,8^. Given the clinical trials findings, and the need to weight the benefits against the risks, vigilant post-marketing surveillance and close monitoring of reported adverse events following vaccination (AEFI) has become indispensable ^5,14^. Driven by public interest and concerns, this study aims to evaluate the post-marketing safety profile of RSV vaccine (RSVPreF) among pregnant individuals in the U.S.

## METHODS

### Data Source

We used the Vaccine Adverse Event Reporting System (VAERS) database, a U.S. national spontaneous-reporting system for monitoring potential vaccine safety signals and identifying adverse events (AEs) that may require further investigation. VAERS gathers reports on post-vaccination AEs, including pregnancy-related complications, from healthcare providers and various stakeholders, with the CDC advocating for reporting any clinically significant AEs affecting mothers or infants ^15,16^.

VAERS captures demographic details, medical history, specifics of the AE, and vaccine-related data. AEs are coded using the Medical Dictionary for Regulatory Activities (MedDRA), with each report assigned various MedDRA Preferred Terms (PTs) that represent signs, symptoms, and diagnostic results, without necessarily confirming a medical diagnosis.

This study followed the Strengthening the Reporting of Observational Studies in Epidemiology (STROBE) reporting guideline ^17^.

### Eligibility Criteria

To examine the safety profile of RSVPreF vaccine during pregnancy following its U.S. approval, we extracted all reports of AEFI from VAERS database spanning from September 1, 2023, to February 23, 2024. To identify AE reports among pregnant individuals within our dataset, we narrowed the scope of extracted reports to those of females age 18 to 49 years, and leveraged MedDRA coding alongside text-string searches, following Moro and colleagues’ methodology ^18^. Our approach included automated searches for reports with MedDRA terms related to pregnancy and perinatal conditions, specific terms indicating exposure during pregnancy, and a text search for “preg” in symptom descriptions, histories, and current illness fields, filtering out any negations of pregnancy (online supplemental table S1). Reports fitting any of these criteria were included in our analysis dataset for further evaluation.

### Statistical Analysis

#### Descriptive Analyses

Descriptive analyses included frequencies of reported AEs, maternal age, gestational age at time of vaccination, interval from vaccination to the onset of AE, reported outcomes, and seriousness of the report.

Given that clinical trial findings where the majority of serious AEs reported by maternal participants vaccinated with the RSVPreF vaccine—irrespective of causality—were related to pregnancy complications such as preterm births and hypertensive disorders (e.g., pre-eclampsia, gestational hypertension) ^8,9,19^, these AEs were considered of special interest, warranting close vigilance. Accordingly, for surveillance purposes, we defined “preterm birth” using the diagnostic codes from the Brighton collaboration – Preterm Birth and Assessment of Gestational Age Companion guide ^20^, which employs specific MedDRA codes to precisely define and identify medical concepts of preterm birth. These terms have been considered relevant to study hypotheses related to preterm birth as a vaccine-product related reaction ^20^. For additional details, refer to the supplementary material (online supplemental table S2).

#### Data Mining (Disproportionality Analyses)

For disproportionality analyses, we employed the Bayesian Confidence Propagation Neural Network (BCPNN) ^21^, chosen for its ability to handle low expected counts and stabilize the observed-to-expected ratios even in data-sparse scenarios ^22^. BCPNN method uses Bayesian statistics within a neural network framework to calculate the Information Component (IC), a logarithmic measure comparing the observed to expected reporting rates of specific vaccine-AE pair under the assumption of no vaccine-AE association ^23^. Utilizing this method, we calculated signal scores for AEs associated with RSVPreF, assessing how specific RSVPreF-AEFI combinations are different from the entire database. This includes comparisons with all spontaneously reported AEFI for other FDA-approved vaccines in pregnant individuals, such as inactivated influenza, COVID-19, and pertussis vaccines. We estimated the IC using the method by Noren and colleagues (2011) ^21^, where a signal is flagged if the 2.5% quantile of the posterior distribution of the IC (IC_025_) exceeds zero, indicating a potential disproportionality that warrants closer examination ^23^.

Adhering to methodologies from previous studies ^18,24^, we excluded reports from our disproportionality analysis where the vaccine type was unspecified or pertained to vaccines contraindicated in pregnancy by the FDA, including live attenuated influenza, MMR, and varicella vaccines ^25^. We also excluded terms not evaluable as suspected adverse reactions, such as MedDRA terms related to pregnancy exposure and diagnostic testing, ensuring consistency with the approaches of comparable studies ^26^.

Descriptive and inferential statistical analyses were conducted using R Statistical Software.

#### Clinical Reviews

Guided by disproportionality analysis, our clinical review, led by D.M., an obstetrics and gynecology physician, focused on RSVPreF AEFI reports with significant disproportionality scores related to pregnancy-specific conditions, primarily focusing on preterm birth cases. The review aimed to validate identified cases, distinguishing between new cases and those with pre-existing conditions or complications, then flagging ineligible cases for exclusion.

#### Patient and public involvement

None.

## RESULTS

Between September 2023 and February 2024, VAERS database received a total of 547 reports that met the study’s criteria for pregnancy related AEFI reports. Of these, 77 reports pertained to RSVPreF vaccination, with maternal age spanning from 21 to 41 years. A significant portion, 76.7%, of vaccinations occurred during the third trimester, with a median onset time of one day for adverse events post-vaccination. Over half of the RSVPreF-linked reports, 54.6%, were deemed serious; among these, 47.5% led to hospitalizations, 25.4% to doctor or clinic visits, and 23.7% to emergency room or urgent care visits. Table 1 provides further details on the characteristics of VAERS reports following RSVPreF vaccination.

**Table 1.** Characteristics of VAERS Reports Received Following RSVPreF Vaccine in Pregnant Individuals, United States, September 1, 2023 - February 23, 2024.

| Characteristic |  |
| --- | --- |
| Total reports | 77 |
| Serious reports, n (%)<sup>a</sup> | 42 (54.55) |
| Maternal age in years, median (range) | 32 (21-41) |
| Gestational age in weeks at time of vaccination, median (range) | 35 (9-38) |
| Interval from vaccination to adverse event in days, median (range) | 1 (0-37) |
| Number of reports given with other vaccines, n (%) | 6 (7.79) |
| Type of facility administering the vaccine, n (%) |  |
| Pharmacy or store | 36 (46.75) |
| Private | 27 (35.06) |
| Unknown | 7 (9.09) |
| Other | 2 (2.60) |
| Public | 2 (2.60) |
| Workplace clinic | 2 (2.60) |
| Military | 1 (0.01) |
| Reported Outcome, n (%)<sup>b</sup> |  |
| Hospitalized | 28 (47.46) |
| Doctor or other healthcare professional office/clinic visit | 15 (25.42) |
| Emergency room/department or urgent care visit | 14 (23.73) |
| Disability | 1 (1.69) |
| Congenital anomaly or birth defect | 1 (1.69) |

<sup>a</sup> Reports are classified as serious in accordance with the Code of Federal Regulations criteria, which classify an AEFI report as serious if it involves death, necessitates hospitalization or extends a current hospital stay, is life-threatening, or leads to a substantial and lasting disability.
<sup>b</sup> A single VAERS report may have multiple outcomes.

Reports submitted to VAERS often included multiple terms to describe the AEs, which can be grouped into three main categories ^27^: non-pregnancy-specific (such as local and systemic reactions), pregnancy-specific, and fetus-related AEs (Table 2). In total, 211 suspected adverse reaction term associated with RSVPreF vaccine were documented, comprising 143 non-pregnancy-specific terms (67.8%), 61 pregnancy-specific AE terms (28.9%), and 7 fetus-related AE terms (3.3%). For non-pregnancy-specific AEs, the most common terms included headache, injection site erythema, and injection site pain, reported at rates of 3.8%, 3.8%, and 2.8%, respectively. Preterm birth emerged as the leading pregnancy-specific AE at 12.8%, with premature separation of placenta, cesarean section, cervical dilatation, pregnancy hemorrhage, and uterine contractions each reported less frequently.

**Table 2.**
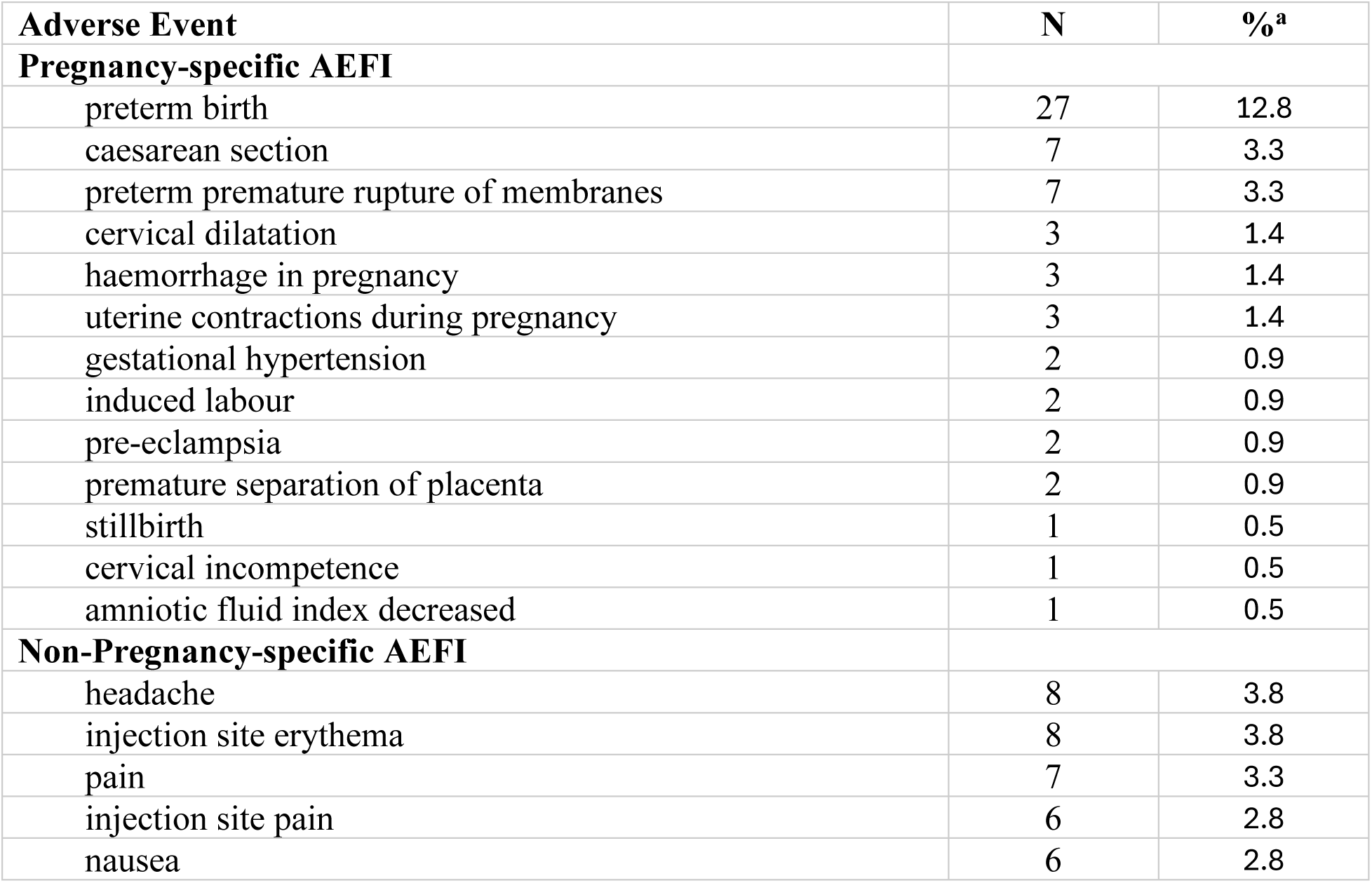
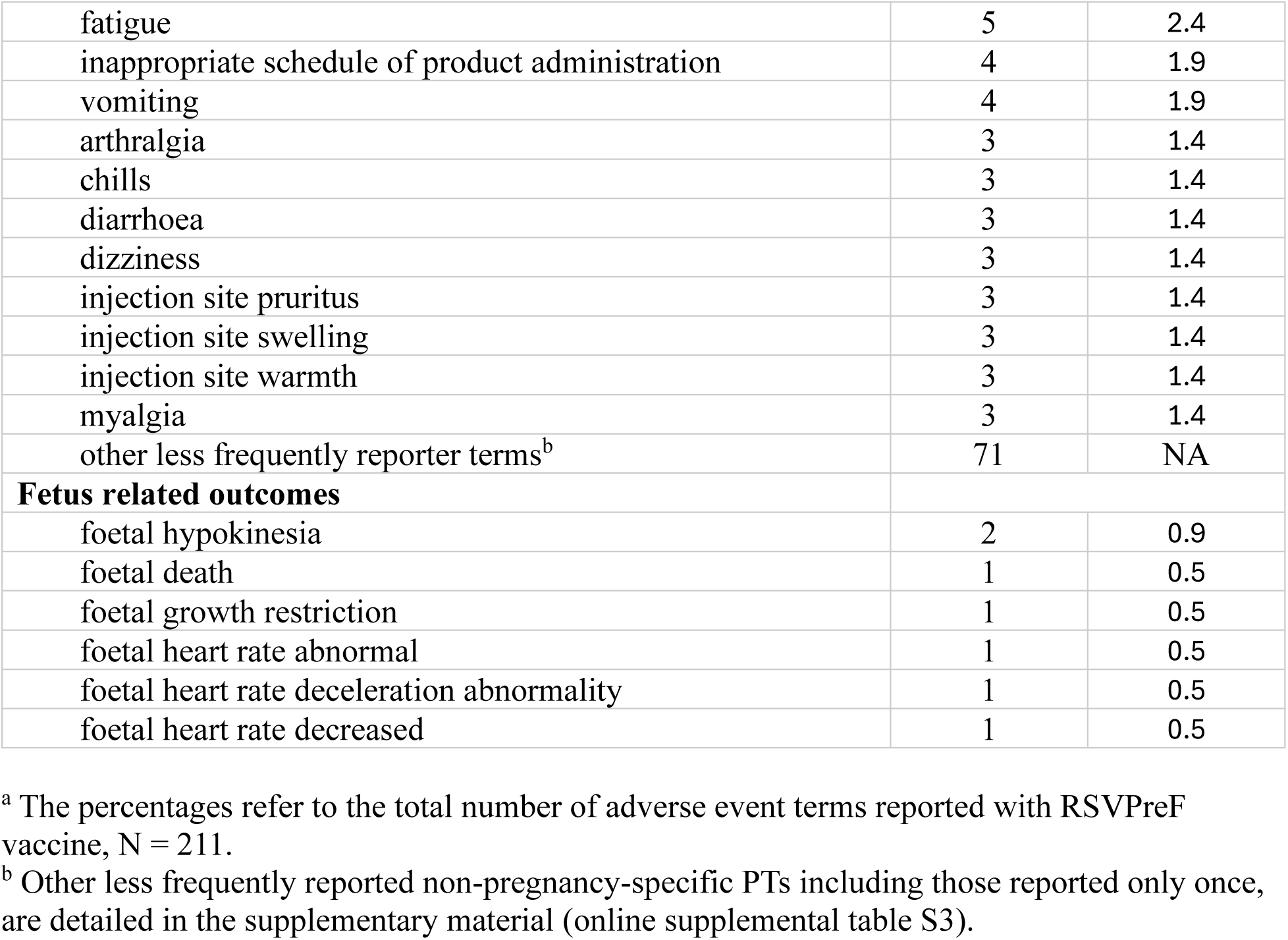
Most Reported Adverse Event Terms Following RSVPreF Vaccination in VAERS.

| **Adverse Event** | **N** | **%<sup>a</sup>** |
| --- | --- | --- |
| **Pregnancy-specific AEFI** |  |  |
| preterm birth | 27 | 12.8 |
| caesarean section | 7 | 3.3 |
| preterm premature rupture of membranes | 7 | 3.3 |
| cervical dilatation | 3 | 1.4 |
| haemorrhage in pregnancy | 3 | 1.4 |
| uterine contractions during pregnancy | 3 | 1.4 |
| gestational hypertension | 2 | 0.9 |
| induced labour | 2 | 0.9 |
| pre-eclampsia | 2 | 0.9 |
| premature separation of placenta | 2 | 0.9 |
| stillbirth | 1 | 0.5 |
| cervical incompetence | 1 | 0.5 |
| amniotic fluid index decreased | 1 | 0.5 |
| **Non-Pregnancy-specific AEFI** |  |  |
| headache | 8 | 3.8 |
| injection site erythema | 8 | 3.8 |
| pain | 7 | 3.3 |
| injection site pain | 6 | 2.8 |
| nausea | 6 | 2.8 |
| fatigue | 5 | 2.4 |
| inappropriate schedule of product administration | 4 | 1.9 |
| vomiting | 4 | 1.9 |
| arthralgia | 3 | 1.4 |
| chills | 3 | 1.4 |
| diarrhoea | 3 | 1.4 |
| dizziness | 3 | 1.4 |
| injection site pruritus | 3 | 1.4 |
| injection site swelling | 3 | 1.4 |
| injection site warmth | 3 | 1.4 |
| myalgia | 3 | 1.4 |
| other less frequently reporter terms<sup>b</sup> | 71 | NA |
| **Fetus related outcomes** |  |  |
| foetal hypokinesia | 2 | 0.9 |
| foetal death | 1 | 0.5 |
| foetal growth restriction | 1 | 0.5 |
| foetal heart rate abnormal | 1 | 0.5 |
| foetal heart rate deceleration abnormality | 1 | 0.5 |
| foetal heart rate decreased | 1 | 0.5 |

<sup>a</sup> The percentages refer to the total number of adverse event terms reported with RSVPreF vaccine, N = 211.
<sup>b</sup> Other less frequently reported non-pregnancy-specific PTs including those reported only once, are detailed in the supplementary material (online supplemental table S3).

### Disproportionality Analysis

In our disproportionality analysis focusing on RSVPreF vaccine AEFI with three or more reports, a signal was detected for preterm birth, marked by statistically significant disproportionality score with an IC value of 2.18 (95% Confidence Interval [CI]: 1.54-2.63). In addition, this analysis revealed significant signals for other MedDRA terms such as caesarean section, preterm premature rupture of membranes, cervical dilatation, injection site pain, warmth, erythema, and inappropriate product administration scheduling, as outlined in online supplemental table S4.

Notably, no signals were identified for haemorrhage in pregnancy, and conditions such as gestational hypertension, stillbirth, and pre-eclampsia were not assessed as they each had fewer than three reports, falling below the evaluation threshold.

### RSVPreF vaccine and preterm birth

In evaluating AEs of special interest associated with RSVPreF vaccination, preterm birth emerged with a significant signal of disproportionate reporting, suggesting that the specific combination of RSVPreF-preterm birth has been reported more frequently than statistically expected when compared with the background of all cases reported to the VAERS database, indicating a potential vaccine safety issue ^23^. The median maternal age of these cases was 33 years, ranging from 25 to 40. According to the WHO classification ^11^, the identified cases were further categorized as follows:

- No cases of extremely preterm (<28 weeks);
- A single case of very preterm (28–<32 weeks);
- Twenty-one cases fell into the moderate or late preterm category (32–<37 completed weeks of gestation); and
- Information on gestational age was not available for five cases.

The median time from RSVPreF vaccination to the onset of preterm delivery was 3 days, spanning from 0 to 31 days, and notably, two-thirds of the cases occurred within a week of immunization (online supplemental table S5). Upon further examination of these preterm birth AEFI reports, instances of co-vaccination were uncommon, only involving two cases—one with the Tdap vaccine and another with the COVID-19 vaccine administered alongside RSVPreF. The majority, 92.6% (25 cases), of these preterm birth reports were deemed serious, with hospitalization being the most frequent outcome for nearly 67.6% (25 cases) of them.

Additionally, 24% (9 cases) required emergency or urgent care visits, two cases led to doctor or clinic visits, and there was a single instance of disability reported. Notably, there were no reports of death or life-threatening events.

In the comprehensive clinical review of these identified preterm birth reports, no cases were excluded, however assessing the certainty level of cases (definite, probable, or possible) presented challenges due to the varying completeness of the reports. Essential information required for this assessment ^20^, such as details of maternal history, date of the last menstrual period or the date of assisted reproductive technology interventions, ultrasound scan results, maternal physical exams, fundal height measurements, newborn birth weight, and physical exams, were often missing. This lack of key data hindered our ability to assess causality and accurately classify the cases with a high degree of certainty.

## DISCUSSION

This study marks the first post-authorization safety analysis of the RSVPreF vaccine among pregnant individuals, utilizing passive surveillance data to characterize the safety profile of the vaccine in a larger and more heterogeneous population than those observed in pre-licensure trials^28^. We leveraged the VAERS database, which can be utilized to perform near real-time surveillance on vaccine safety. Nonetheless, VAERS is not designed to assess causal associations; its primary purpose is to detect potential vaccine safety concerns that may warrant further investigations ^15^.

The safety profile of RSVPreF in pregnant individuals, previously assessed in clinical studies ^12^ ^29^, highlighted self-limiting AEs like injection site pain and headaches, with a slight increase in pre-eclampsia rates among the vaccine group ^30^. However, concerns about preterm births emerged ^8,14^, underscoring the need for ongoing surveillance, particularly given the trial’s exclusion of individuals at higher risk for preterm delivery. According to the trial protocol, the manufacturer was examining preterm birth as an AE of special interest ^12,14^. However, pregnant individuals at higher risk for preterm delivery, due to factors such as high BMI, IVF pregnancies, alongside other risk factors, were excluded from both phase 2b and phase 3 trials of RSVPreF vaccine ^8,19^. Similarly, those with a history of pregnancy complications were generally not enrolled ^19^.

In our analysis, the pattern of reported non-pregnancy-specific AEs mirrored those identified during RSVPreF vaccine’s pre-licensure phase. However, among pregnancy-specific AEs, preterm birth emerged as the most reported AEFI, with cesarean section, preterm premature rupture of membranes, cervical dilatation, pregnancy hemorrhage, and uterine contractions also observed. A closer look at preterm birth cases showed a median time of 3 days from vaccination to AE onset, with two-thirds occurring within the first week after immunization. This finding particularly diverges from clinical trial safety outcomes, where the majority of preterm births were reported more than 30 days post-vaccination ^8^. Moreover, our disproportionality analysis highlighted a significant safety signal for preterm birth, indicating a stronger association between the RSVPreF vaccine and reporting of preterm birth as an AEFI than what would be expected by chance. Nevertheless, the inherent limitations of the information provided in VAERS reports preclude a definitive establishment of a causal relationship between RSVPreF vaccination and preterm birth.

Understanding the complex and largely unknown pathophysiology behind preterm birth is essential, particularly given the various maternal, fetal, and placental factors at play ^31^. This complexity in causation underscores the importance of approaching these vaccine-related AEs through a wider comprehensive lens. When we compare the pregnancy-specific symptomatology associated with RSVPreF vaccine with those associated with other vaccines recommended during pregnancy, distinct differences in the pregnancy-specific AE profiles can be noticed. For seasonal influenza vaccines, spontaneous abortion (fetal death occurring < 20 weeks gestation) was the most commonly reported pregnancy-specific AE, followed by stillbirth (fetal death occurring ≥ 20 weeks gestation), with 6 reported preterm birth cases, accounting for only 1.1% of reported AEs ^24^. The 2009 H1N1 influenza vaccine similarly had miscarriage as the leading reported AE, with reports also noting stillbirths, and 7 cases of preterm births constituting just 2.4% of reported AEFI ^32^. For Hepatitis A and Hepatitis AB vaccines, spontaneous abortions were the most frequently reported AEs, with preterm deliveries being less common (5.0% or 7 cases) alongside elective terminations ^18^. With the COVID-19 vaccine, spontaneous abortion was the most commonly pregnancy-related AEs reported to VAERS, with preterm delivery being comparatively rare (0.9% or 2 cases) ^27,33^. These patterns drawn from the VAERS database for various vaccines and across different time intervals, diverge with the reported AE profile following RSVPreF vaccination, where preterm birth was the most frequently pregnancy-specific AE reported, underscoring a unique AE profile for the RSVPreF vaccine in pregnant individuals.

The strengths of our study stem from leveraging VAERS, a comprehensive pharmacovigilance system, with a broad national scope, capacity for near-real time surveillance, and adeptness at detecting rare AEFI ^34^. However, it’s essential to interpret our findings within the context of VAERS’ intrinsic limitations. Our analysis, reliant on participant-reported data, is limited by the lack of comprehensive information on several risk factors for adverse pregnancy outcomes, such as complete maternal history, lifestyle behaviors (including smoking and drug use), comorbidities, and infections ^35^. In addition, VAERS is prone to various reporting biases ^36^, including over-reporting, stimulated reporting, and under-reporting, which despite mandatory reporting requirements, are probably substantial for pregnancy- and neonatal-specific AEs ^27^.

Also, events temporally close to vaccination are more likely to being reported ^24^; however, reporting is still dependent on and influenced by medical suspicion, which can be influenced by the perception of a causal relationship with the vaccine, even among health care providers who underreport events that are not clearly related to vaccination ^37^. Notable limitations also include our inability to ascertain the total number of RSVPreF vaccine doses administered, and the absence of an unvaccinated comparator group, which restricts our ability to estimate incidence rates and relative risks from VAERS data alone. Although we detected a signal for preterm birth linked to the RSVPreF vaccine, establishing causality from numerical data alone is challenging. Therefore, our findings should be considered as hypothesis-generating, necessitating further research.

## CONCLUSION

Findings from this passive pharmacovigilance study indicate a safety signal for preterm birth among pregnant individuals vaccinated with RSVPreF vaccine. Despite the inherent constraints of passive surveillance systems, our analysis contributes to the current understanding of the safety of this vaccine in pregnancy. Further research, potentially via pregnancy registries or by leveraging existing healthcare databases for prospective cohort studies is imperative to further explore this potential safety signal.

## Data Availability

All data produced are available online at

https://vaers.hhs.gov/data/datasets.html

## DATA SHARING STATEMENT

All the data generated or analyzed during this study are publicly available on the Vaccine Adverse Events Reporting System (VAERS) and are available for download from: https://vaers.hhs.gov/data/datasets.html

## AUTHOR CONTRIBUTIONS

AA and DM designed the study. AA handled the acquisition, analysis, and interpretation of data. AA drafted the manuscript and conducted the statistical analysis. DM provided clinical assessment. SP-L and DM critically reviewed and revised the manuscript. All authors approved the final version for publication. AA is responsible for the overall content as guarantor. DM supervised the study.

## CONFLICT OF INTEREST DISCLOSURES

The authors report no conflict of interest.

## FUNDING

This research received no specific grant from any funding agency in the public, commercial or not-for-profit sectors.

## PATIENT AND PUBLIC INVOLVEMENT

Patients and/or the public were not involved in the design, or conduct, or reporting, or dissemination plans of this research.

## PATIENT CONSENT FOR PUBLICATION

Not applicable.

## ETHICS APPROVAL

This study utilizes publicly available, de-identified data and is based on previously published documents. It does not involve human or animal subjects; therefore, ethical approval is not required.

## Notes

### Competing Interest Statement

The authors have declared no competing interest.

