## Supplementary material for "Safety of RSV Vaccine among Pregnant Individuals: A Real-World Pharmacovigilance Study Using Vaccine Adverse Event Reporting System": Online supplemental table S1, Online supplemental table S2, Online supplemental table S3, Online supplemental table S4, Online supplemental table S5

This supplementary material aims to provide comprehensive insights into the research methodology and additional information about the results of the analysis.

**Online supplemental table S1.** Search Strategy to Identify Cases of Adverse Events Following Immunization Among Pregnant Individuals.

**Online supplemental table S2.** MedDRA Terms For Preterm Birth Case Definition and Identification.

**Online supplemental table S3.** Reported Adverse Events in Pregnant Women Following Receipt of RSVPreF Vaccine in Pregnant Individuals, VAERS, September 2023 – February 2024.

**Online supplemental table S4.** Disproportionality Analysis of Adverse Events Reported with RSV Vaccines Compared to All Other Vaccines in Pregnant Individuals, VAERS, September 2023 – February 2024.

**Online supplemental table S5.** Distribution of Identified Preterm Birth Cases by the Time Interval Between RSVPreF Vaccination and Reported Preterm Delivery.

**Supplementary Material References.**

### Online supplemental table S1. Search Strategy to Identify Cases of Adverse Events Following Immunization Among Pregnant Individuals

To Identify adverse events following immunization among pregnant individuals, the following automated search strategy was utilized, following Moro et al. (2014)'s methodology[1]:

| **Search strategy:** | |
|---|---|
| i) | reports that included MedDRA terms within the two system organ classes (SOC), “Pregnancy, Puerperium, and Perinatal Conditions” and “Congenital, Familial, and Genetic Disorders”; |
| ii) | reports that included the following MedDRA terms: “Maternal exposure timing unspecified”, “Maternal exposure during pregnancy”, “Exposure during pregnancy”, “Paternal exposure during pregnancy”, “Foetal exposure during pregnancy”; and |
| iii) | a text string search for the term “preg” within each report to query fields such as symptom descriptions and adverse event narratives (symptom_text), pre-existing conditions (history), and current illnesses (cur_ill), with instructions to exclude mentions of ‘not pregnant’, ‘non-pregnant’, ‘non pregnant’, ‘nonpregnant’, and ‘no preg’. |
| Reports that met any of these delineated criteria were included in our analysis dataset for further evaluation. | |

### Online supplemental table S2. MedDRA Terms For Preterm Birth Case Definition and Identification

| LLT code | LLT name | PT code | PT name |
|---|---|---|---|
| 10015873 | Extreme immaturity | 10036590 | Premature baby |
| 10014051 | Early onset of delivery, unspecified as to episode of care | 10036595 | Premature delivery |
| 10062694 | Premature baby less than 26 weeks | 10036590 | Premature baby |
| 10053593 | Premature baby 26 to 32 weeks | 10036590 | Premature baby |
| 10053594 | Premature baby 33 to 36 weeks | 10036590 | Premature baby |
| 10004953 | Birth premature | 10036590 | Premature baby |
| 10014049 | Early onset of delivery | 10036595 | Premature delivery |
| 10036594 | Premature birth | 10036590 | Premature baby |
| 10003969 | Baby premature | 10036590 | Premature baby |
| 10021410 | Immature baby | 10036590 | Premature baby |
| 10021734 | Infant premature | 10036590 | Premature baby |
| 10036615 | Prematurity | 10036590 | Premature baby |
| 10075861 | Preterm labour | 10036600 | Premature labor |
| 10023555 | Labour premature | 10036601 | Premature labor |

| | | | |
|---|---|---|---|
| 100140 49 | Early onset of delivery | 100365 95 | Premature delivery |
| 100767 29 | Very preterm infant | 100365 90 | Premature baby |
| 100766 88 | Late preterm infant | 100365 90 | Premature baby |
| 100324 05 | Other preterm infants | 100365 90 | Premature baby |
| 100140 50 | Early onset of delivery, delivered, with or without mention of antepartum condition | 100365 95 | Premature delivery |
| 100039 67 | Baby 28 weeks plus under 2.5 kg | 100365 90 | Premature baby |
| 100324 15 | Other preterm infants, unspecified weight | 100365 90 | Premature baby |
| 100324 14 | Other preterm infants, less than 500 grams | 100365 90 | Premature baby |
| 100049 53 | Birth premature | 100365 90 | Premature baby |

**LLT Code:** This column contains the unique identifier assigned to a "Lowest Level Term" (LLT) within the MedDRA hierarchy. LLTs represent the most specific level of terminology and are used to capture the precise details of an adverse event or medical history term reported in clinical and post-marketing settings.

**LLT Name:** This column specifies the name or description of the Lowest Level Term (LLT). These names are detailed and specific, reflecting the exact term as reported or documented in a clinical or regulatory context.

**PT Code:** This column contains the unique code assigned to a "Preferred Term" (PT) in MedDRA. Preferred Terms are a higher-level classification within the MedDRA hierarchy, under which one or more LLTs are grouped.

**PT Name:** In this column, the standardized name of the Preferred Term (PT) is provided.

### Online supplemental table S3. Reported Adverse Events in Pregnant Women Following Receipt of RSVPreF Vaccine in Pregnant Individuals, VAERS, September 2023 – February 2024

| MedDRA term | N |
|---|---|
| abdominal discomfort | 1 |
| abdominal pain upper | 1 |
| accidental exposure to product | 1 |
| administration site erythema | 1 |
| amniotic fluid index decreased | 1 |
| arthralgia | 3 |
| back pain | 1 |
| blood iron decreased | 1 |
| blood pressure increased | 2 |
| brain fog | 1 |
| caesarean section | 7 |
| cervical dilatation | 3 |
| cervical incompetence | 1 |
| chills | 3 |
| complication of pregnancy | 1 |
| cough | 1 |
| decreased appetite | 1 |
| diarrhoea | 3 |
| dizziness | 3 |
| dyskinesia | 1 |
| dyspnoea | 1 |
| erythema | 1 |
| eyelid function disorder | 1 |
| facial asymmetry | 1 |
| facial pain | 1 |
| facial paralysis | 1 |
| fatigue | 5 |
| feeling hot | 1 |
| foetal death | 1 |
| foetal growth restriction | 1 |
| foetal heart rate abnormal | 1 |
| foetal heart rate deceleration abnormality | 1 |
| foetal heart rate decreased | 1 |
| foetal hypokinesia | 2 |
| gestational hypertension | 2 |
| haemorrhage in pregnancy | 3 |
| hand-foot-and-mouth disease | 1 |
| headache | 8 |

| | |
|---|---|
| heart rate increased | 1 |
| herpes zoster | 1 |
| high risk pregnancy | 1 |
| hypertension | 2 |
| inappropriate schedule of product administration | 4 |
| incorrect route of product administration | 1 |
| induced labour | 2 |
| influenza | 1 |
| injection site erythema | 8 |
| injection site hypersensitivity | 1 |
| injection site induration | 1 |
| injection site infection | 1 |
| injection site mass | 2 |
| injection site pain | 6 |
| injection site pruritus | 3 |
| injection site reaction | 1 |
| injection site swelling | 3 |
| injection site warmth | 3 |
| liver function test increased | 1 |
| loss of consciousness | 1 |
| muscle spasms | 1 |
| muscular weakness | 1 |
| musculoskeletal stiffness | 1 |
| myalgia | 3 |
| nasal congestion | 1 |
| nasopharyngitis | 1 |
| nausea | 6 |
| neck pain | 1 |
| night sweats | 1 |
| oropharyngeal pain | 1 |
| pain | 7 |
| pain in extremity | 1 |
| pallor | 1 |
| palpitations | 1 |
| peripheral swelling | 2 |
| placental calcification | 1 |
| precipitate labour | 1 |
| pre-eclampsia | 2 |
| premature separation of placenta | 2 |
| preterm premature rupture of membranes | 7 |
| preterm birth | 27 |
| protein urine present | 2 |
| pyrexia | 2 |

| | |
|---|---|
| rash | 2 |
| respiratory tract congestion | 1 |
| rhinorrhoea | 2 |
| sciatica | 1 |
| sensitive skin | 1 |
| stillbirth | 1 |
| swelling | 2 |
| urine protein/creatinine ratio increased | 1 |
| urticaria | 1 |
| uterine contractions during pregnancy | 3 |
| uterine hypertonus | 1 |
| vaginal discharge | 1 |
| vaginal haemorrhage | 1 |
| viral infection | 1 |
| visual impairment | 1 |
| vomiting | 4 |

### Online supplemental table S4. Disproportionality Analysis of Adverse Events Reported with RSV Vaccines Compared to All Other Vaccines in Pregnant Individuals, VAERS, September 2023 – February 2024

| Adverse Event | N | IC ($IC_{025}$, $IC_{975}$) |
|---|---|---|
| preterm birth | 27 | 2.18 (1.54, 2.63) |
| headache | 8 | 0.90 (-0.31, 1.70) |
| injection site erythema | 8 | 1.52 (0.30, 2.32) |
| caesarean section | 7 | 1.65 (0.34, 2.50) |
| pain | 7 | 0.32 (-0.98,1.18) |
| preterm premature rupture of membranes | 7 | 2.06 (0.76, 2.91) |
| injection site pain | 6 | 1.42 (0.01, 2.34) |
| nausea | 6 | 0.76 (-0.65, 1.67) |
| fatigue | 5 | 0.50 (-1.07, 1.48) |
| inappropriate schedule of product administration | 4 | 1.84 (0.07, 2.92) |
| vomiting | 4 | 0.62 (-1.15, 1.70) |
| arthralgia | 3 | 0.20 (-1.87, 1.41) |
| cervical dilatation | 3 | 2.42 (0.35, 3.63) |
| chills | 3 | 0.69 (-1.38, 1.89) |
| diarrhoea | 3 | 1.20 (-0.87, 2.41) |
| dizziness | 3 | 1.69 (-0.38, 2.89) |
| haemorrhage in pregnancy | 3 | 2.01 (-0.06, 3.22) |
| injection site pruritus | 3 | 2.01 (-0.06, 3.22) |
| injection site swelling | 3 | 1.42 (-0.65, 2.63) |
| injection site warmth | 3 | 2.42 (0.35, 3.63) |
| myalgia | 3 | 0.69 (-1.38, 1.89) |
| uterine contractions during pregnancy | 3 | 1.20 (-0.87, 2.41) |

IC: Information Component; $IC_{025}$ and $IC_{975}$: Lower and Upper Bounds of the 95% Credibility Interval for IC.

Numbers in red indicated a disproportionality signal was detected.

### Online supplemental table S5. Distribution of Identified Preterm Birth Cases by the Time Interval Between RSVPreF Vaccination and Reported Preterm Delivery.

| Interval | Number of cases |
|---|---|
| <1 week after immunisation | 18 |
| 1–<2 weeks after immunisation | 4 |
| 2–<4 weeks after immunisation | 3 |
| 4–<6 weeks after immunisation | 2 |
